# Assortative mating across the full spectrum of mental disorders: a nationwide Finnish register study

**DOI:** 10.1101/2025.04.28.25326538

**Authors:** Kateryna Golovina, Mai Gutvilig, Ripsa Niemi, Christian Hakulinen

## Abstract

**Background:** Previous research has shown assortative mating across various psychiatric disorders; however, their definitions of partnership have often been limited, and the timing of relationship formation remains imprecise. This study aims to comprehensively examine assortative mating across the full spectrum of mental disorders, using population-wide register data from Finland that include information on the formation of both marriages and cohabiting unions.

**Methods:** We used nationwide data on all cohabitations and marriages between 2000 and 2020 from the Finnish Population Register. Broad and specific categories of mental disorder diagnoses were retrieved from both primary and secondary healthcare registers in Finland. We calculated tetrachoric correlations between partners’ mental disorder diagnoses, considering only diagnoses received before the start of cohabitation or marriage.

**Results:** The study included 964,017 men and 957,207 women, forming 1,271,242 partnerships. Assortative mating was observed across the full spectrum of mental disorders, with the strongest within-disorder correlations for schizophrenia, psychotic disorders, organic mental disorders, and intellectual disabilities (r > 0.50). Moderate correlations were found for mood and anxiety disorders. Adjusting for birth decade and excluding comorbidities slightly attenuated the associations but did not change the overall patterns.

**Conclusions:** This study suggests that assortative mating is prevalent for mental disorders. These findings suggest that assortative mating may contribute to the transmission and clustering of mental disorders within families, highlighting the importance of considering partner selection in mental health research and policy-making.

## Introduction

Assortative mating, or non-random mating, refers to individuals’ tendency to choose partners with similar traits. This phenomenon has been documented across various characteristics (1), including mental and somatic health (2). A range of mental disorders has been examined, such as attention-deficit/hyperactivity disorder (ADHD), bipolar disorders, autism spectrum disorder (ASD), depression, and substance use disorders, but previous studies were mainly based on cohort samples with healthy volunteer selection bias (3–6). To date, only two studies relied on population-wide register-based data from Sweden and Norway and explored a broader range of mental disorders (7,8). The strongest within-disorder correlations were found for ADHD, ASD, schizophrenia, and substance use disorders. The between-disorder correlations were also widespread, but the strengths of associations varied (7,8). These studies provided evidence for direct and indirect assortative mating, where partners resemble each other due to selection either on the observed trait or a correlated trait (8).

However, the definition of partnership varies across studies, and it might not always be possible to disentangle convergence (i.e., whereby a disorder may develop in one partner due to the influence of the other) from assortative mating. For example, a study from Sweden defined partnership as either marriage or biological parenthood, without requiring the disorder to be diagnosed prior to partnership formation (7). A Norwegian study anchored partnership to parenthood and deduced the beginning of the relationship based on the time of first childbirth (8). These definitions of partnership, which rely heavily on couples with children, may introduce selection bias, as prior research has shown that people with mental disorders are less likely to become parents (9). Moreover, there is evidence that spousal convergence is common for certain mental disorders, such as substance use disorders (10). Therefore, it is important to distinguish between convergence and assortative mating.

In the present study, we comprehensively examine assortative mating across the full spectrum of mental disorders using both primary and secondary healthcare register data from Finland. To overcome the limitations of previous research, we apply a broader definition of partnership, including all unions based on cohabitation and marriage. To focus solely on the assortative mating process, we include only those people who were diagnosed with a mental disorder prior to the start of their cohabitation/marriage. Our first objective is to evaluate the magnitude and significance of partner correlations within and between all mental disorders in a population-based sample. Our second objective is to examine the strength of these associations after controlling for birth decade, and comorbidities. Moreover, as previous studies primarily relied on couples with children (7,8), we also compared our results to analyses limited to partners with shared children enabling us to assess whether the associations differ when these restrictions are applied.

## Methods and Materials

### Study population

Data on partnerships, as well as on birth year and any shared children, were retrieved from the full Finnish Population Register (FOLK) of Statistics Finland. The dataset includes the beginning and end dates of cohabitations, marriages, and registered partnerships. Cohabitations were defined according to the definition of Statistics Finland as two individuals of different sexes and over the age of 18 who live in the same residence, are no more than 16 years apart in age, and are not siblings (11). However, if a couple shares children, there criteria do not apply (11). All individuals who got married or started cohabitating at some point between January 1^st^, 2000, and December 31^st^, 2020, were included in the study population. The study population was restricted to different-sex partnerships based on each individual’s registered gender as of June 2024 as the data does not cover all same-sex marriages, registered partnerships, or cohabitations. If a couple cohabited/married more than once, the formation date of the first partnership was used. If a person had multiple partnerships, all partnerships were included.

Partnership data was subsequently linked to data on mental disorders diagnosed prior to the formation of the partnership. Mental disorder diagnoses were retrieved from the Care Register for Health Care (CRHC) and the Register of Primary Healthcare Visits (RPHV) of the Finnish Institute for Health and Welfare. Individual-level register linkages were conducted using personal pseudonymized identity numbers, which are assigned to all Finnish residents. The ethics committee of the Finnish Institute for Health and Welfare approved the study plan (THL/184/6.02.01/2023§933). Data were linked with the permission of Statistics Finland (TK-53-1696-16) and the Finnish Institute for Health and Welfare. Informed consent is not required for register-based studies in Finland.

### Measures

Mental disorder diagnoses (International Classification of Diseases 10th revision [ICD-10] F10 to F99 and equivalent ICD-8, ICD-9, and International Classification of Primary Care, 2nd edition [ICPC-2] codes), included inpatient hospital episodes (1970-2020, secondary outpatient visits (1998–2020), and primary healthcare visits (2011–2020) in Finland. In our study, broad categories of mental disorders across the whole ICD-10 subchapter F were used: organic mental disorders, including dementia (F00-F09), substance use disorders (F10-F19), psychotic disorders (F20-F29), mood disorders (F30-F39), anxiety disorders (F40-F48), behavioral syndromes associated with physiological disturbances and physical factors (F50-F59), personality disorders (F60-F69), intellectual disabilities (F70-F79), developmental disorders (F80-F89), and childhood onset disorders (F90-F98). We also used several specific categories of mental disorders: schizophrenia (F20), bipolar disorder (F30-F31), depressive disorder (F32-F34), eating disorders (F50), and sleep disorders (F51), pervasive developmental disorders (F84), and hyperkinetic disorders (F90). Finally, we also examined any mental disorder (F00-F99) as its own category. A person was considered diagnosed if they received their diagnosis prior to the start date of the cohabitation/marriage and undiagnosed if they had no record of a diagnosis or the first diagnosis occurred only after the start of cohabitation/marriage.

### Statistical analysis

Following the approach in Torvik et al. (8), we calculated the prevalence of each mental disorder by whether an individual’s partner had also been diagnosed with the same disorder prior to the start of marriage or cohabitation. Next, we calculated tetrachoric correlations both within and across disorders to evaluate the magnitude and significance of the association between partners’ diagnostic status. To account for potential cohort effects, we calculated partial correlations adjusted for both individuals’ birth decades. To evaluate whether the partner correlations differ in partners who share children and those who do not, we stratified the analyses by whether the couple had children together at any point.

We also conducted two sensitivity analyses to evaluate the robustness of the findings. First, as the relationship formation tends to happen prior to cohabitation/marriage, we repeated the main analysis with only those diagnosed at least five years prior to cohabitation/marriage considered diagnosed. Second, following the study by Nordsletten et al. (7) and to reduce confounding from comorbidities, we repeated the main analysis using only the main ICD-10 subchapter F categories. In this analysis, we excluded women diagnosed with the male partners’ disorder of interest and men diagnosed with the female partners’ disorder of interest. For example, when examining correlations between male mood disorders and female anxiety disorders, this analysis included only men without anxiety disorders and women without mood disorders.

For all correlations, we present findings from analyses where the expected contingency table cell frequencies were at least five or greater for all cells and observed cell frequencies were greater than zero. Where applicable, p-values were corrected for multiple comparisons using the Bonferroni method, with significance set at 0.05.

Data management was performed using Stata version 17.0 (12). Statistical analyses, figures, and tables were generated in R version 4.2.2 (13) using the packages *lavaan*, *polycor*, and *tidyverse* (14–16).

## Results

The study population included 964,017 men and 957,207 women, forming 1,271,242 partnerships. **Table 1** shows the prevalence of mental disorder diagnoses among men and women. Before the start of cohabitation/marriage, 15.6% of men and 19.7% of women received a mental disorder diagnosis. Anxiety disorders were the most common in the study population, followed by mood disorders and substance use disorders in men, and behavioral and emotional syndromes in women. On average, men entered cohabitation or marriage at age 28 (interquartile range [IQR] = 23–37), and women at age 26 (IQR = 21–34). The median age at diagnosis for each mental disorder category is shown in **Supplementary Table 1**. Overall, 39.1% of the partnerships had children.

**Table 1.** Prevalence of mental disorder diagnoses among men and women in the study population.

| **Diagnosis** | **Men, % (n)** | **Women, % (n)** |
| --- | --- | --- |
| Any Mental Disorder (F00-F99) | 15.56% (197782) | 19.66% (249868) |
| Organic Mental Disorders (F00-F09) | 0.41% (5210) | 0.35% (4509) |
| Substance Use Disorders (F10-F19) | 4.74% (60248) | 3.24% (41221) |
| Psychotic Disorders (F20-F29) | 0.99% (12534) | 1.18% (15001) |
| Schizophrenia (F20) | 0.3% (3845) | 0.34% (4298) |
| Mood Disorders (F30-F39) | 5.19% (65977) | 9.52% (121079) |
| Bipolar Disorder (F30-F31) | 0.65% (8287) | 1.06% (13473) |
| Depressive Disorder (F32-F34) | 4.96% (63072) | 9.35% (118808) |
| Anxiety Disorders (F40-F48) | 6.37% (81024) | 10.25% (130240) |
| Behavioural & Emotional Syndromes (F50-F59) | 1.84% (23382) | 3.75% (47682) |
| Eating Disorders (F50) | 0.1% (1214) | 1.51% (19231) |
| Sleep Disorders (F51) | 1.55% (19739) | 2.22% (28183) |
| Personality Disorders (F60-F69) | 1.63% (20691) | 1.81% (23030) |
| Intellectual Disabilities (F70-F79) | 0.21% (2672) | 0.21% (2704) |
| Developmental Disorders (F80-F89) | 1.47% (18733) | 1.03% (13032) |
| Pervasive Developmental Disorders (F84) | 0.13% (1699) | 0.08% (1060) |
| Childhood Onset Disorders (F90-F98) | 2.39% (30418) | 2.6% (33082) |
| Hyperkinetic Disorder (F90) | 0.81% (10350) | 0.42% (5311) |

Figure 1 shows the prevalence of mental disorder diagnoses among men and women with unaffected and affected partners. People with affected partners were more likely to have the same diagnosis themselves, although the strength of this pattern varied by diagnosis. The greatest differences were observed for organic mental disorders (e.g., dementia), substance use disorders, schizophrenia, and intellectual disabilities.

**Figure 1.**
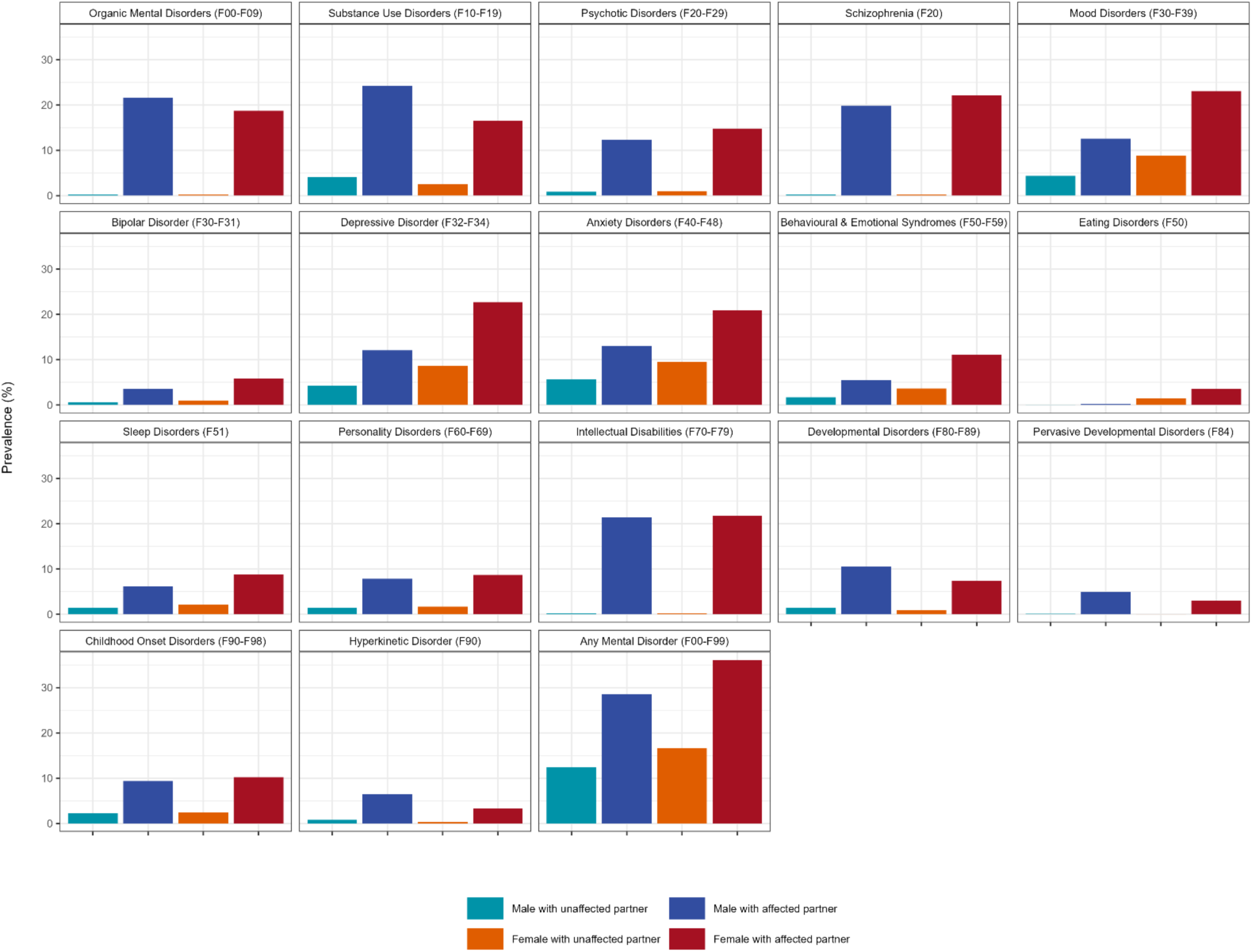
Prevalence of mental disorders among men and women with unaffected and affected partners.

Figure 2 shows a heatmap of tetrachoric correlations within and between male and female partners’ mental disorder diagnoses. The strongest within-disorder correlations (>0.50) were found for organic mental disorders, psychotic disorders, schizophrenia, and intellectual disabilities. In contrast, within-disorder correlations for more common mental disorders (e.g., mood disorders, anxiety disorders, and bipolar disorders) ranged from 0.20 to 0.40, with the weakest correlation found for eating disorders (r = 0.11).

**Figure 2.**
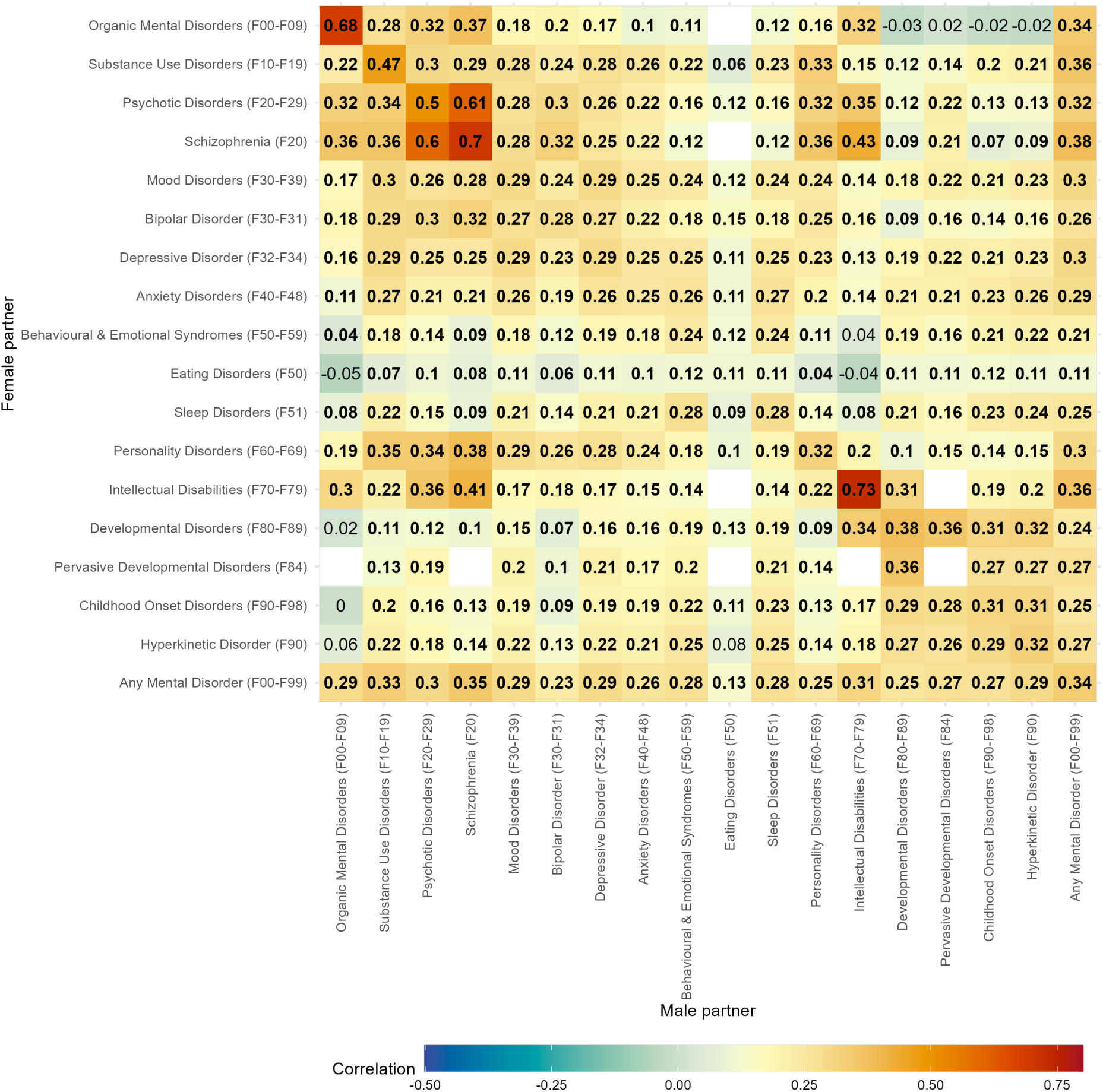
Tetrachoric correlations within and between mental diagnoses of partners. Bold figures indicate a statistically significant correlation (Bonferroni corrected p < 0.05). Empty values reflect the contingency table had an observed cell frequency of 0 or an expected cell frequency of less than 5.

The strongest between-disorder correlations were observed primarily among severe and early-onset disorders, particularly psychotic and neurodevelopmental conditions (Figure 2). Psychotic and schizophrenia disorders showed the highest cross-partner associations: female partner’s psychotic disorder and male partner’s schizophrenia (r = 0.61), and vice versa – male psychotic disorder and female schizophrenia (r = 0.60); Female schizophrenia and male intellectual disabilities (r = 0.43), and male schizophrenia and female intellectual disabilities (r = 0.41). Personality disorders in female partners were moderately correlated with a range of male partner diagnoses, including, schizophrenia (r = 0.38), substance use disorder (r = 0.35), male psychotic disorder (r = 0.34), mood disorders (r = 0.29), bipolar disorder (r = 0.28), and anxiety disorder (r = 0.24). Similar patterns were observed in the opposite direction – male personality disorders were also linked to female partners’ schizophrenia, substance use, and other disorders. Finally, developmental and childhood-onset disorders were also associated across partners. Female developmental disorders were related to male partners’ pervasive developmental disorders, intellectual disabilities, hyperkinetic disorder, and other childhood-onset conditions. These associations were slighly weaker in the reverse direction, but overall indicating similar patterns for male developmental disorders and female partners’ diagnoses.

These associations slightly attenuated after adjusting for birth decade (**Supplementary Figure 1**). Similar patterns were observed among partners without children (**Supplementary Figure 2A**) and when considering only diagnoses made more than five years before cohabitation (**Supplementary Figure 3**). Among partners with children, many associations are not shown due to insufficient data, and the observed associations were slightly attenuated compared to the main model (**Supplementary Figure 2B**). For instance, the associations between female partners’ personality disorder and male partners’ schizophrenia, substance use disorders, and psychotic disorders were reduced compared to the primary analysis. A similar pattern was observed in the opposite direction, that is, male partners’ personality disorders and the corresponding disorders in female partners.

After excluding comorbidities, the strongest associations were observed between organic mental disorders and intellectual disabilities (r = 0.29), organic mental disorders and psychotic disorders (r = 0.27), and psychotic disorders and intellectual disabilities (r = 0.25). See **Supplementary Figure 4** for details.

## Discussion

This study provides a systematic examination of assortative mating across the full spectrum of mental disorders using nationwide register data from Finland. Our findings align with previous research from Sweden and Norway (7,8), demonstrating strong assortative mating for schizophrenia, intellectual disabilities, psychotic disorders, organic mental disorders, and substance use disorders. Additionally, we observed widespread between-disorder correlations among partners with varying strengths. The adjustments for birth decade, having children, comorbidities, and diagnosis made five years before cohabitation slightly attenuated the associations, but did not change them.

The strong within-disorder correlations for severe mental disorders may suggest that men and women with these diagnoses are particularly likely to form partnerships with others who have the same disorder. This may be explained by shared genetic vulnerabilities, environmental exposures, or social homogamy, where individuals with similar life circumstances and experiences are more likely to meet and form relationships (2). Indirect assortative mating may also explain some of the findings, as recently shown (8). Partners may not assort directly on a specific mental disorder but rather on correlated traits, such as personality characteristics or cognitive functioning, which in turn increase the likelihood of shared diagnoses. For other mental disorders, including mood and anxiety disorders, assortative mating was present but to a lesser extent. These weaker correlations may reflect the higher prevalence of these disorders in the general population and their heterogeneity in symptoms, making assortative mating less pronounced. Nevertheless, significant partner correlations indicate that selection into relationships based on mental health status is not random.

Our sensitivity analyses showed that adjusting for birth decade slightly attenuated associations but did not change our findings, indicating that cohort effects do not fully explain partner correlations. When restricting the analyses to diagnoses made more than five years before cohabitation, similar patterns were observed, suggesting that assortative mating occurs prior to partnership formation rather than being driven by shared environmental influences during the early stages of the relationship prior to cohabitation/marriage. Among partners with children, some associations were not presented due to insufficient data, and associations were slightly attenuated. This attenuation could be due to selection effects where individuals with severe mental disorders are less likely to have children (9,17,18). However, the general trend of assortative mating remained and is in line with previous register-based studies from Sweden and Norway which largely relied on couples with children (7,8). Additionally, after excluding comorbidities, the strongest cross-disorder associations were between female partners’ organic mental disorders and male partners’ intellectual disabilities, organic mental disorders and psychotic disorders, as well as female partners’ psychotic disorders and male partners’ intellectual disabilities. These associations were mirrored in the opposite direction – male partners’ organic mental disorders or psychotic disorders and the corresponding disorders in female partners.

## Strengths and limitations

The strength of our study was the use of both primary and secondary health care data from the Finnish nationwide registers, enabling us to examine the associations with high statistical accuracy and minimal health-related selection biases. Register-based mental disorder diagnoses are generally shown to have good validity, but some conditions lack proper validation, and variations may occur across clinical settings (19). Additionally, our broader definition of partnership allowed a more comprehensive examination of assortative mating, as the population register includes complete data on all marriages, cohabitations, and registered partnerships from 2000 to 2020.

However, the present findings should be interpreted in light of several limitations. First, although our data included individuals who were treated in both primary and secondary healthcare settings, those who did not seek help or who were treated in primary care before year 2011 were not considered. Additionally, the data lack information on the onset, remission, and recovery of mental disorders. Therefore, we cannot distinguish whether individuals formed relationships, for example, during the active phases of the disorder or after recovery. Second, we did not have the information on the date of relationship formation, only the date of marriage, cohabitation, or registered relationship. To eliminate this limitation, we conducted sensitivity analysis by including only those diagnosed at least five years prior to cohabitation, marriage, and registered relationship. Finally, our study was restricted to different-sex partnerships, as same-sex relationships were not systematically recorded in the data. Future studies should examine whether similar assortative mating patterns hold in same-sex couples.

## Conclusion

In conclusion, this study found that assortative mating is prevalent both within and across the full spectrum of mental disorders. The strongest associations were found for severe mental disorders, including schizophrenia, psychotic disorders, organic mental disorders, and intellectual disabilities. Common mental disorders, such as mood and anxiety disorders, showed moderate correlations with each other but also frequently co-occurred with other mental disorders, especially severe ones. While these findings are largely in line with previous research (7,8), our subgroup analyses suggest that earlier findings may be attenuated due to relying on partners with shared children. Taken together, these findings support the role of assortative mating as one of the mechanisms contributing to the transmission and clustering of mental disorders within families.

## Acknowledgements

This study was supported by the Otto A. Malm Foundation (KG); the European Union’s Horizon 2022 Research and Innovation Programme (KG, grant number 101094741); the Research Council of Finland (CH, grant number 354237); and the European Union (ERC, MENTALNET, grant number 101040247 to CH). The proposal for this work was reviewed by their dedicated peer reviewers and assessment panel prior to funding being awarded. The funder otherwise played no role in this research. Views and opinions expressed are those of the authors only and do not necessarily reflect those of the European Union or the European Research Council. Neither the European Union nor the granting authority can be held responsible for them.

## Data availability statement

The data used in this study are the property of Statistics Finland and the Finnish Institute of Health and Welfare. The data are available from the respective authorities, but certain restrictions apply. For more information on accessing the data, please visit www.stat.fi.

## Details of Ethics Approval

The ethics committee of the Finnish Institute for Health and Welfare approved the study plan (THL/184/6.02.01/2023§933). Data were linked with the permission of Statistics Finland (TK-53-1696-16) and the Finnish Institute for Health and Welfare. Informed consent is not required for register-based studies in Finland.

## Supplementary materials

**Supplementary Table 1.** Median Age at Diagnosis.

| Diagnosis | Men, Median<br>(IQR) | Women, Median<br>(IQR) |
| --- | --- | --- |
| Any Mental Disorder (F00-F99) | 21 (16–30) | 20 (16–28) |
| Organic Mental Disorders (F00-F09) | 51 (26–76) | 64 (30–80) |
| Substance Use Disorders (F10-F19) | 26 (20–36) | 22 (18–33) |
| Psychotic Disorders (F20-F29) | 25 (20–33) | 24 (19–31) |
| Schizophrenia (F20) | 26 (22–33) | 26 (21–34) |
| Mood Disorders (F30-F39) | 26 (20–36) | 22 (17–31) |
| Bipolar Disorder (F30-F31) | 30 (23–39) | 25 (20–34) |
| Depressive Disorder (F32-F34) | 25 (20–36) | 22 (17–31) |
| Anxiety Disorders (F40-F48) | 22 (19–30) | 22 (17–29) |
| Behavioral & Emotional Syndromes (F50-F59) | 24 (19–34) | 19 (16–26) |
| Eating Disorders (F50) | 17 (13–21) | 17 (15–21) |
| Sleep Disorders (F51) | 24 (19–34) | 22 (18–31) |
| Personality Disorders (F60-F69) | 23 (19–31) | 24 (20–31) |
| Intellectual Disabilities (F70-F79) | 16 (8–25) | 16.5 (9–27) |
| Developmental Disorders (F80-F89) | 9 (6–12) | 10 (6–15) |
| Pervasive Developmental Disorders (F84) | 12 (8–16) | 14 (10–19) |
| Childhood Onset Disorders (F90-F98) | 12 (8–15) | 14 (11–16) |
| Hyperkinetic Disorder (F90) | 13 (8–19) | 18 (13–25) |

**Supplementary Figure 1.**
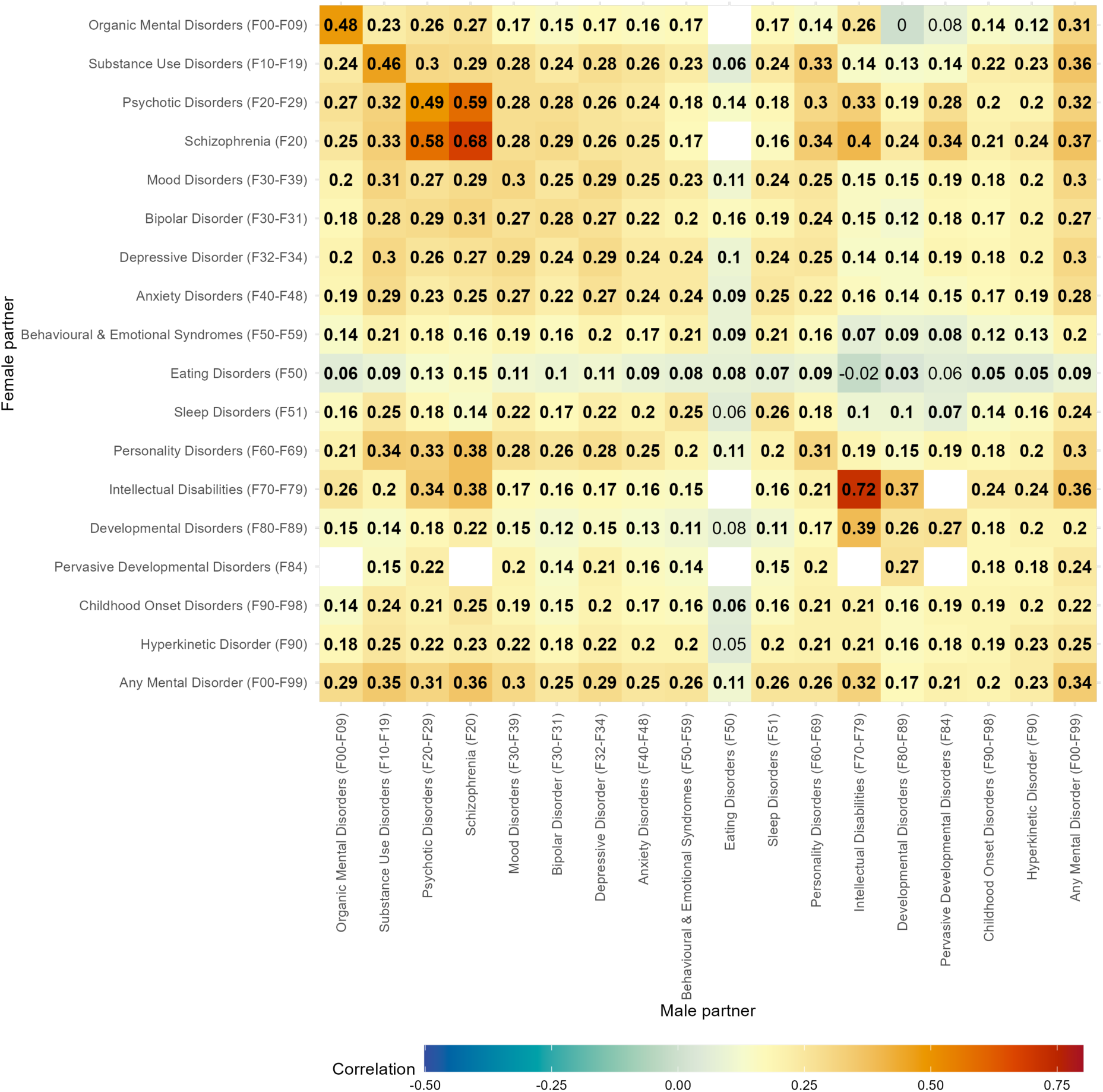
Tetrachoric correlations within and between mental diagnoses of partners adjusted for a birth decade.

**Supplementary Figure 2A.**
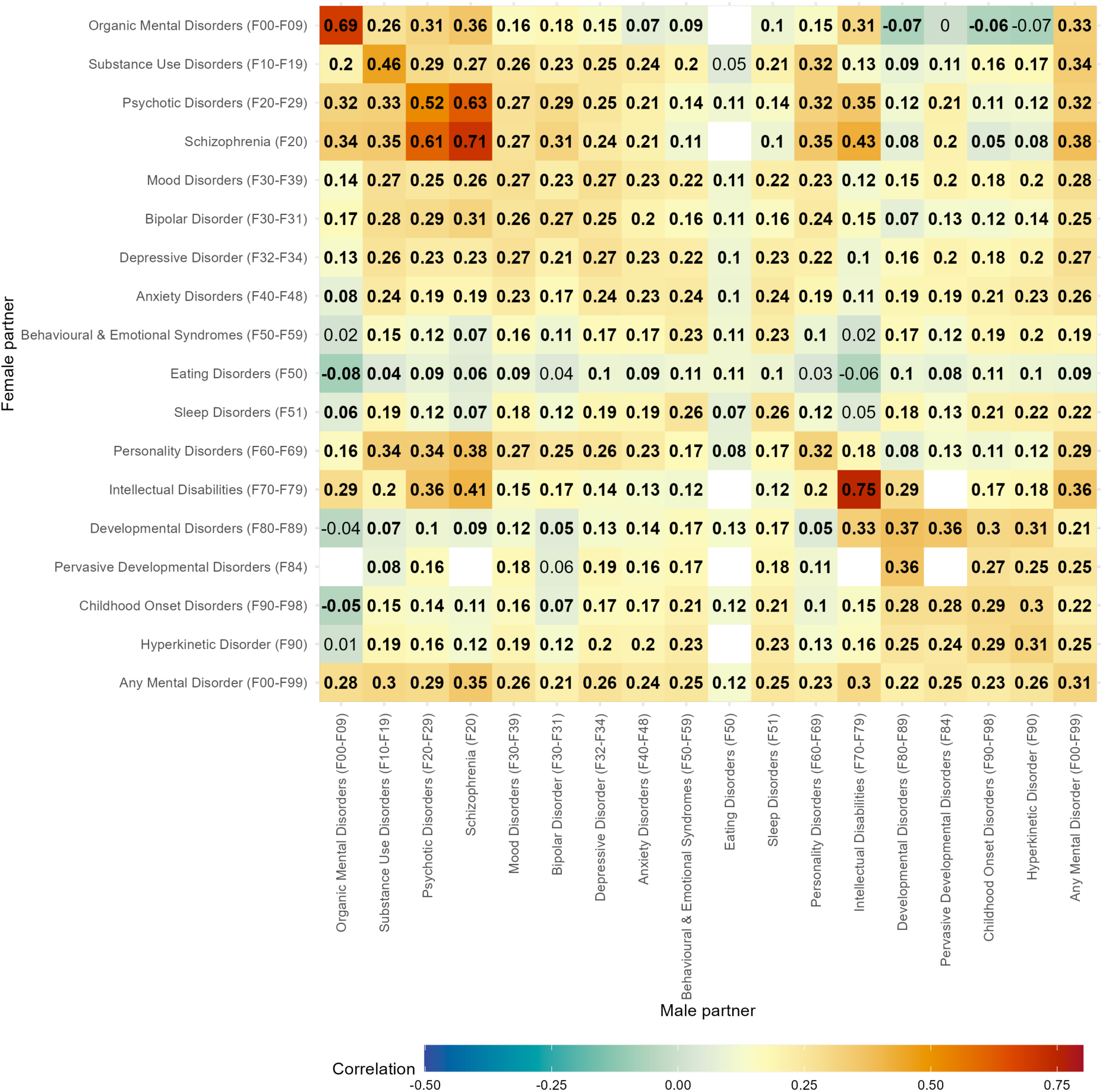
Tetrachoric correlations within and between mental diagnoses of partners without children.

**Supplementary Figure 2B.**
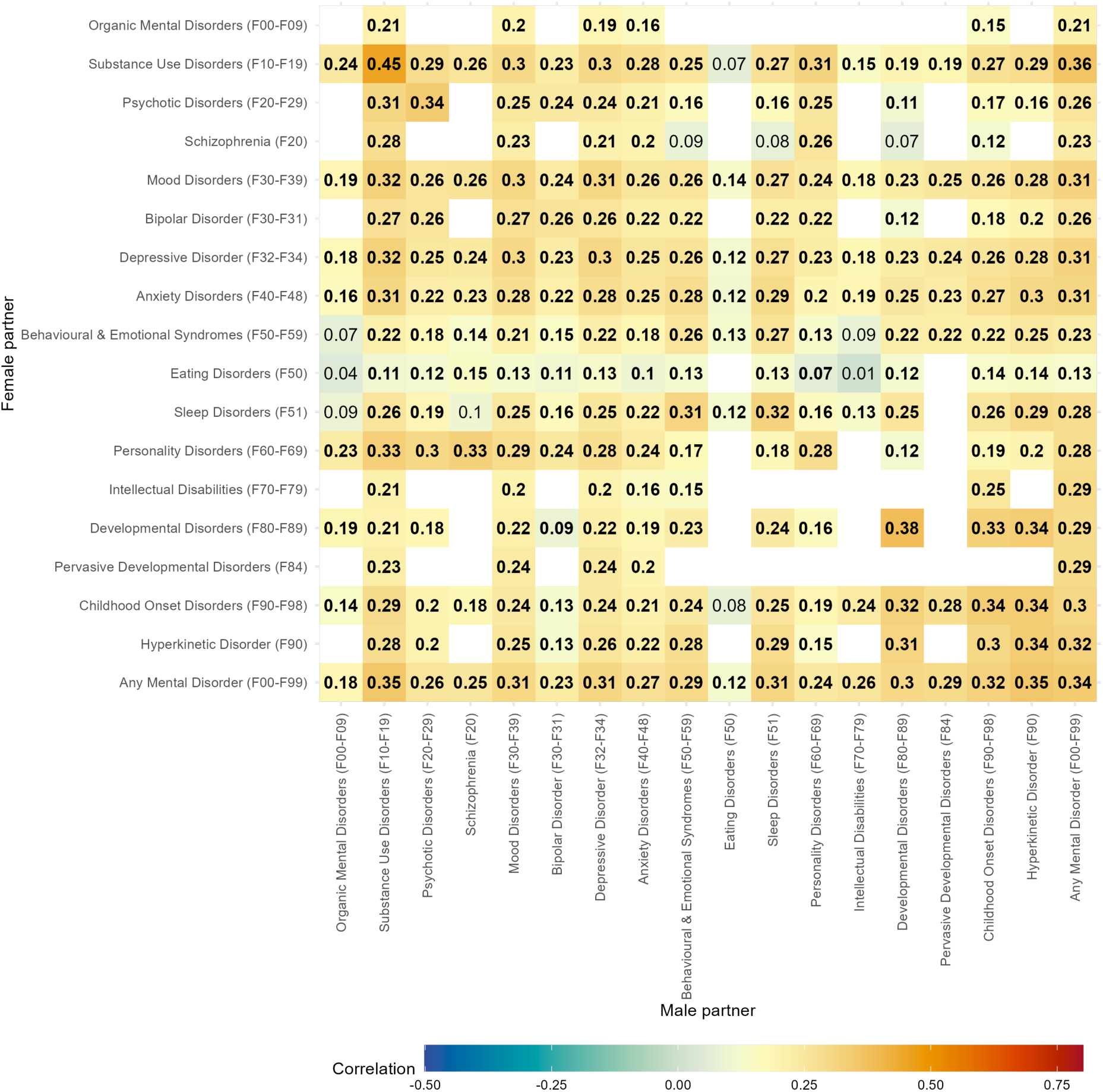
Tetrachoric correlations within and between mental diagnoses of partners with children.

**Supplementary Figure 3.**
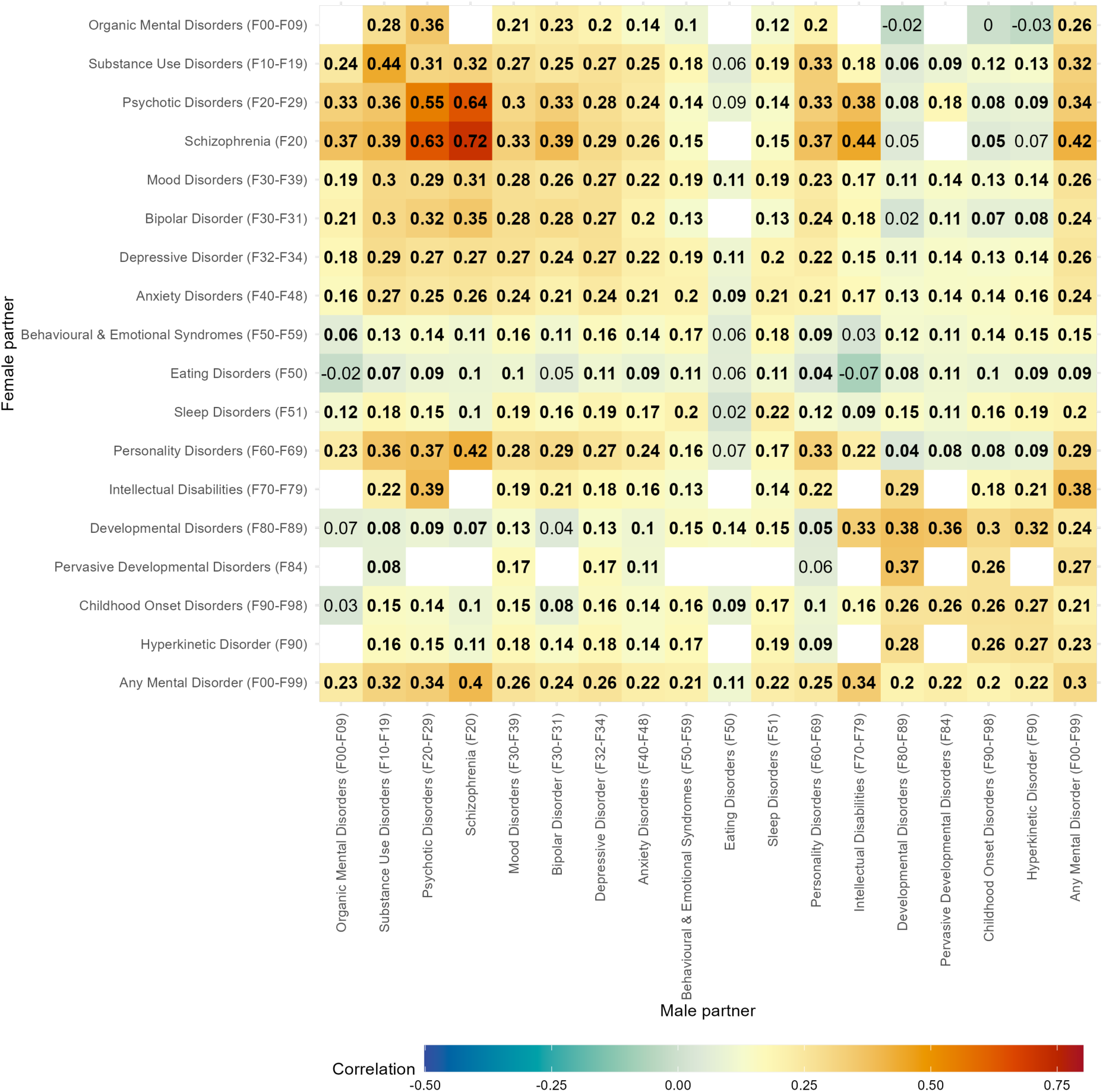
Tetrachoric correlations within and between mental diagnoses of partners including only diagnoses 5 years before cohabitation.

**Supplementary Figure 4.**
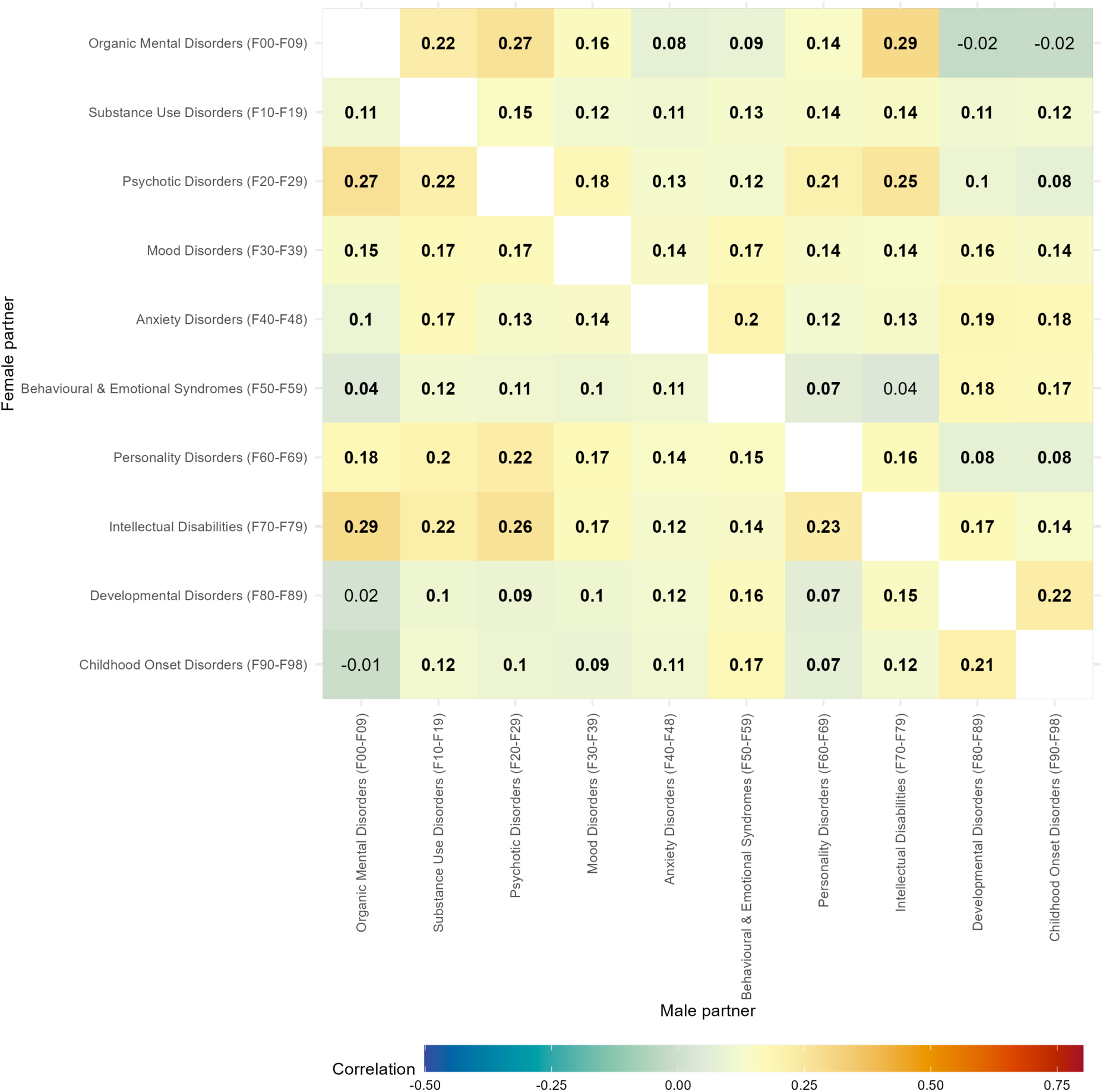
Tetrachoric correlations within and between mental diagnoses of partners when comorbidities were excluded. *Note*. Empty cells indicate that within-disorder analyses could not be conducted because all women diagnosed with the male partners’ disorder of interest were excluded and all men diagnosed with the female partners’ disorder of interest were also excluded.

